# Conservative Management of Acute Appendicitis In The Era Of COVID 19: A Multicenter prospective observational study at The United Arab Emirates

**DOI:** 10.1101/2020.09.30.20204503

**Authors:** Fatima Y. AL Hashmi, Abeer Al Zuabi, Ibrahim Yaseen Hachim, Guido H.H. Mannaerts, Omar Bekdache

**Affiliations:** General surgery Division, Sheikh Shakhbout Medical City, Abu Dhabi, UAE; General surgery Division, Tawam Hospital, Al Ain, UAE; Clinical Sciences Department, College of Medicine, University of Sharjah, UAE; Sharjah Institute for Medical Research, University of Sharjah, UAE; Chief of General Surgery Division, Tawam Hospital, Al Ain, UAE; Director of Trauma Service, Tawam Hospital, Al Ain, UAE

**Keywords:** Acute appendicitis, conservative management, COVID-19

## Abstract

**Background:** Since its emergence in December 2019, the Novel Coronavirus (COVID-19) pandemic resulted in a profound impact on the health care system worldwide. We propose herein to evaluate the impact of implementing conservative management as an alternative approach to surgical appendectomy in the treatment of proven acute appendicitis during COVID19 pandemic.

**Methods:** Our study is a prospective multicenter study that includes a cohort of 160 patients admitted to the surgical departments in both Tawam Hospital and Sheikh Shakhbout Medical City, Abu Dhabi, UAE, for the period from February 2020 till July 2020.

**Results:** Our results showed that 56 of our patients (35%) were treated conservatively, while the other 104 (65%) underwent operative management. There was a significant decrease in length of hospital stay (LOS) (2.32± 0.83 days) among the first group compared to the second (2.8± 1.47 days). Also, short term follow-up showed that 90% of those patients did not require further operative intervention or developed any serious complications. Out of the 110 patients that were swapped for COVID19, nine (8.18%) were confirmed to be positive. Our protocol was to avoid surgical management for COVID19 positive patients unless indicated. This resulted in (8/9) of COVID19 positive patients to be treated conservatively. Follow up was achieved by using telemedicine-based follow-up with the aim of empowering social distancing and reducing risk of viral exposure to patients as well as the health care providers. In conclusion, our results showed that the implementation of conservative management in treating patients with acute appendicitis who were COVID19 positive is a safe and feasible approach that maybe essential in reducing viral transmission risks as well as avoiding operative risks on COVID19 positive patients.

## Introduction

The pandemic of COVID-19 and its associated massive inflow of patients affected many aspects of life with special impact on the health care system(1). The overload of hospitals with COVID-19 patients resulted in changing priorities for the benefit of patients as well as health care providers (2). Also, due to the nature of transmission of the SARS-CoV-2 (COVID-19) that was found to spread during aerosol generating procedures (AGPs) including surgeries, many concerns were raised on infection control and safety on patients as well as health care providers’ (3, 4). For that reason, changes in the management protocols and implementations of new guidelines along with novel practices were recommended with the aim of reducing the risk of infection as well as establishing more efficient use of the available resources. One of the strategies proposed to relieve the burden on the health care system is to avoid hospitalization and defer other non-urgent surgeries.

Acute appendicitis represents one of the most common cause of surgical emergencies and acute abdomen with a lifetime prevalence reaching to 7%(5). The golden standard management plan for such patients is surgical appendectomy (6). However, emerging evidence showed that the conservative non-surgical alternative management by antibiotic therapy might have same impact on the patient’s outcome(6–9). Such approach might have additional advantages compared to the surgical approach including the omission of surgery associated mortality and morbidity including pain, infections and bowel obstruction as well as risks and complications associated with anesthesia and hospitalization(10).

Implementation of conservative management in acute appendicitis may represent an optimal option during the COVID19 pandemic due to its advantages in reducing the length of hospital stay (LOS) and avoiding surgical management with all its risks(11). In addition, patients diagnosed with novel Coronavirus Disease 2019 (COVID-19) may also benefit from such approach due to the fact that they are more vulnerable to the complications owing to their compromised lung functions as well as suppressed immunity and multiple organ dysfunction(12). Moreover, such approach might impact the safety of the staff and health care providers through reduction of their exposure to COVID19 patients.

Here we investigate the effects of implementing conservative approach for the management of acute appendicitis on patient’s outcomes including, length of hospital stay (LOS), duration ofantibiotics use, cost, complications and recurrence, during the COVID-19 outbreak.

### Patients and methods

Our study is a prospective multicenter study that included recruitment of patients admitted to the surgical departments in two tertiary referral university hospitals (Tawam Hospital and Sheikh Shakhbout Medical City) in Abu Dhabi, the capital of the United Arab Emirates, for the period from February 2020 till July 2020. All the epidemiological, clinical as well as the laboratory data of the patients were retracted from Abu Dhabi Health Services Co. (SEHA)electronic file system. This research obtained ethical approval from Abu Dhabi COVID19 Research IRB Committee with reference no. DOH/NCVDC/2020/1045. Our study was performed according to the Strengthening the Reporting of Observational studies in Epidemiology (STROBE) guidelines for observational studies.

### Statistical analysis

Descriptive patients’ data including demographic and laboratory parameters were tabulated and presented as mean and standard deviation in the case of continuous variables. In contrast, frequencies and percentages were used in case of categorical variables. SPSS 26.0 (IBM Corporation, Armonk, NY) software was used for statistical analysis. A p-value of <0.05 was used as a cut-off value to differentiate between significant and non-significant differences. Chi-squared or Fisher’s exact test, in addition to student’s t-test as well as analysis of variance (ANOVA) test were used whenever needed.

## Results

### Patients demographic features

Our cohort consists of 160 patients. The mean age of our patients was 29.61± 10.33, with the majority of them being male patients (63.12%) and female patients representing (36.87%). The mean duration of hospitalization was 2.7± 1.42 days. In addition to clinical evaluation, CT scan was the most common imaging method to confirm the diagnosis (71.15%), followed by US (19.23%). Other demographic and clinical features are described in table 1.

**Table 1.** Comparison of the demographic and clinical parameters of our patient’s cohort consist of 160 patients.

|  | Total | % | Operative management (104) |  | Conservative management (n=56) |  | P |
| --- | --- | --- | --- | --- | --- | --- | --- |
|  |  |  | No. | % | No. | % |  |
| Age (years), mean ±SD | 29.61±10.33 |  | 30.62±11.89 |  | 29.05±9.38 |  | 0.397 |
| Gender |  |  |  |  |  |  |  |
| Male | 101 | 63.12% | 62 | 58.49% | 39 | 72.22% | 0.088 |
| Female | 59 | 36.87% | 44 | 41.50% | 15 | 27.78% |  |
| Imaging |  |  |  |  |  |  |  |
| US | 30 | 19.23% | 20 | 20% | 10 | 17.85% |  |
| CT scan | 111 | 71.15% | 69 | 69% | 42 | 75% |  |
| None | 15 | 9.61% | 11 | 11% | 4 | 7.14% |  |
| Size of appendix (mm), mean ±SD | 10.58±3.51 |  | 11.14± 3.82 |  | 9.78± 2.79 |  | 0.045 |
| Presence of Appendicolith | 34 | 25.95 | 26 | 32.09% | 8 | 16% | 0.04 |
| Absence of Appendicolith | 97 | 74.04 | 55 | 67.90 | 42 | 84% |  |
| Duration of hospitalization (days) , mean ±SD | 2.7±1.42 |  | 2.8±1.47 |  | 2.32±0.83 |  | 0.01 |
| COVID19 negative | 101 | 91.81 | 55 | 98.21% | 46 | 85.18% | 0.031 |
| COVID19 positive | 9 | 8.18 | 1 | 1.78% | 8 | 14.81% |  |
| In patient antibiotic duration , mean ±SD | 1.69±1.3 |  | 1.08±0.63 |  | 2.80±1.48 |  | 8.98503e-12 |
| Total antibiotic duration, mean ±SD | 4.07±4.2 |  | 1.08±0.63 |  | 9.60±1.65 |  | <0.000001 |
| Coast (UAE dirhams), mean ±SD | 9335±4517 |  | 9423±5812 |  | 5804±6254 |  | 0.02 |

### Management option

Out of the 160 patients, 56 patients (35%) were treated conservatively, the other 104 (65%) underwent surgical management. Analysis of the demographic features of both groups revealed no significant difference in the age and sex distribution between both groups (Table 1). Worth mentioning that the group with initial conservative treatment was shown to have smaller size of the appendix (9.78± 2.79mm) compared to the surgical management group (11.14± 3.82) (P= 0.045). Appendicolith was confirmed to be present in 32.09% of the surgical managed group compared to only 16% in the conservatively managed group (P=0.04). As expected, patients with conservative management plan had a significant decrease in length of hospital stay (LOS) (2.32± 0.83 days) compared to patients that underwent surgical management (2.8± 1.47 days). In contrast, the conservatively managed group had longer duration of antibiotic coverage that reached in total to 9.6± 1.65 days compared to only 1.08± 0.63 days for patients who were surgically managed (P=<0.000001). Moreover, the mean total cost per patient for the conservative approach was significantly lower (5804± 6254 United Arab Emirates Dirham (AED) compared to (9423± 5812 AED) for the surgical approach.

### COVID19 status

During the initial phase of COVID19 pandemic (period from February-March, 2020), only 2% of the patients were swabbed for COVID-19. This rate increased to 99.12% in the period from April 2020 onward (Figure1). In total, out of the 160 patients, COVID19 PCR swab test was performed for 110 patients. Nine of those patients (8.18%) were confirmed to be COVID19 positive and the other 101 (91.81%) were confirmed to be negative. Our policy was to avoid surgical management for COVID19 positive patients unless indicated. For that reason, the majority (8/9) of patients that were confirmed to be COVID19 positive, were treated conservatively (Table 1).

**Figure 1:**
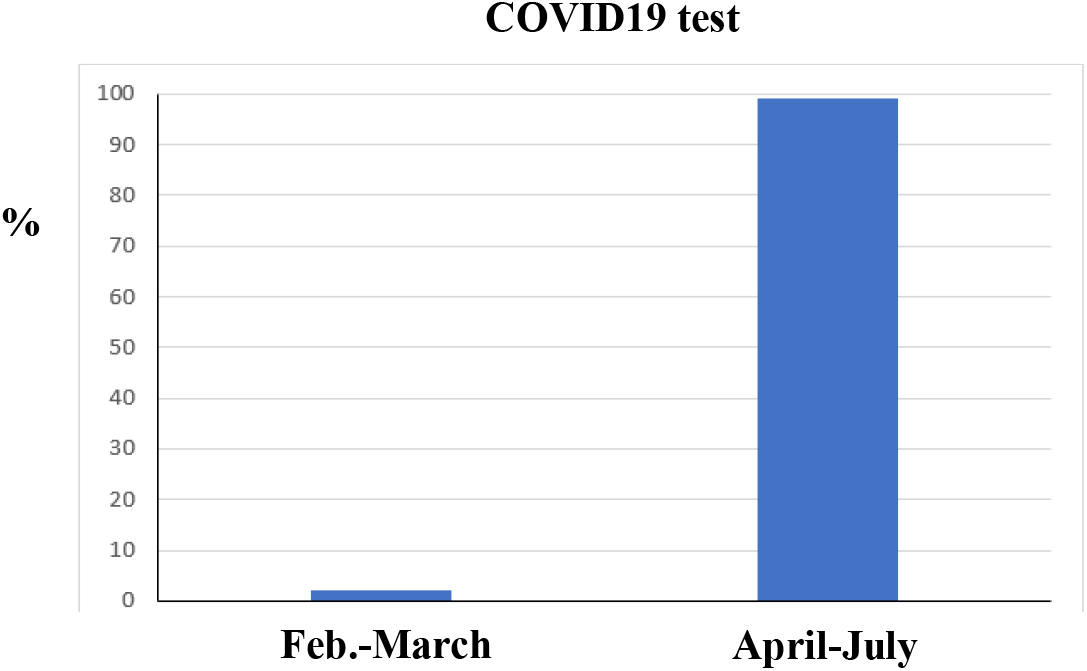
The percentage of COVID19 test for patients in the time period from February to July.

### Patients discharge and short outcome

Monitoring of patients from the time of admission until discharge revealed that both groups of patients showed a similar pattern of improvement that was evident in the improvement of the clinical exam as well as laboratory profile (Table 2). Among the inflammatory markers, our results showed that white blood count (WBC) count was a more sensitive parameter in predicting the clinical improvement compared to the C-reactive protein (CRP) (Table 2). Indeed, WCC was found to be reduced in the operative group from 12.425± 4.636 at time of admission to 8.542± 2.553 at the time of patient’s discharge (P=0.00006). Similarly, the WCC was also found to be significantly reduced in the conservatively treated group from 11.929± 4.313 to 5.331± 1.516 at time of discharge (P=9.32587e-15).

**Table 2.**
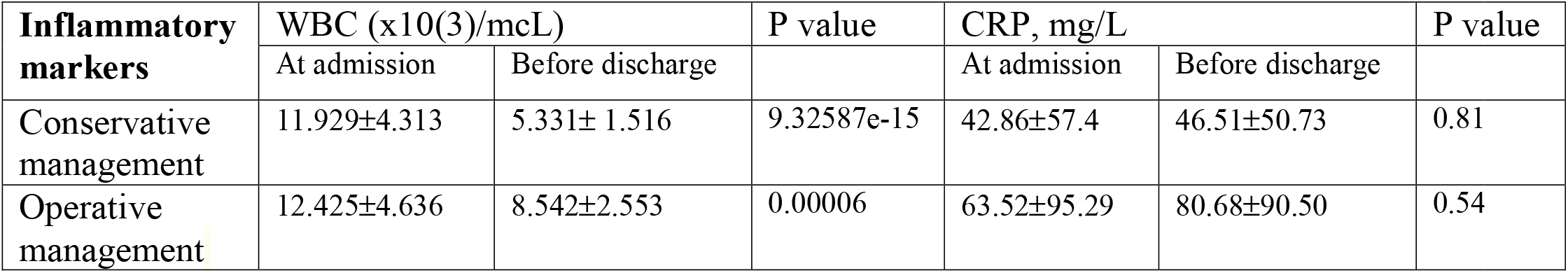
The change in inflammatory markers of conservative and operative treated groups at admission and at discharge.

### Patients follow up and outcome

As shown in table 3, follow up of the patients who received conservative management revealed that only 3 patients (5.35%) return to the ED department within the first week. We were able to follow 28 patients out of 56 (50%) from the same group within 6 months from discharge through either clinic visits or Tele-consultation. Among those, only 3 patients were found to subsequently underwent a surgical appendectomy (10.82%), while the other 25/28 (89.28%) stated no recurrence of symptoms. Importantly, remote follow-up through Tele-consultation was performed in 19 patients out of the 28 cases that were followed up (Figure 2).

**Table 3.** The follow up and outcome of conservative treated group.

| Conservative management | Return to ED within 1 week |  | Follow-up within 6 months including phone follow-up |  |
| --- | --- | --- | --- | --- |
|  | Yes | No | No recurrence/ pain resolved | Appendicectomy |
|  | 3 (5.35%) | 53(94.64%) | 25 (89.28%) | 3 (10.82%) |

**Figure 2:**
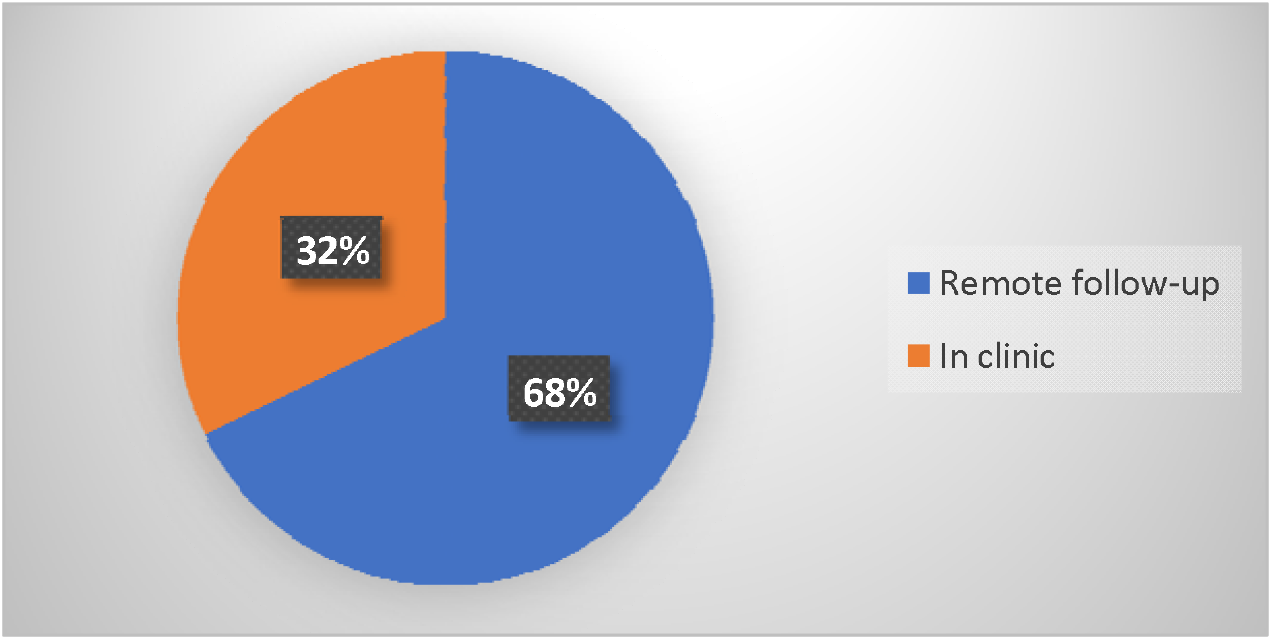
Method of patient’s follow-up.

## Discussion

Since its emergence in December 2019, COVID-19 pandemic resulted in a profound impact on the health systems worldwide(13). The impact of this disease on surgical practices was also significant. This includes its effect on the surgical workforce and hospitals’ infrastructures(14).

Many actions were implemented to improve our response to the disease pandemic with the aim of protecting our patients as well as our health-care providers in addition to increasing hospitals bed capacity to insure a more dynamic use of the available resources(15). One of the main actions taken was surgical prioritization with delaying and deferring hospitalization of non-urgent procedures(15). For that reason, we investigated the effects of implementing conservative approach and compare it to the classical surgical approach in the treatment of confirmed acute appendicitis and its impact on patient’s outcome during COVID-19 pandemic. Our results showed that the implementation of the non-surgical approach resulted in a significant improvement in the clinical presentation and the inflammatory profile of patients, in addition to the reduction in the length of hospital stay (LOS)indicating high success rate. Furthermore, short term follow up of those patients showed that the majority (90%) did not need further operative intervention or develop serious complications. Indeed, this correlates well with other reports implementing conservative management in acute appendicitis to be associated with shorter hospital stay and a low risk of late recurrence(16–18). This clearly demonstrated that the conservative approach for the management of acute appendicitis represents a feasible, safe and effective alternative to the surgical approach. In addition, our finding revealed that the mean cost of conservative management was around half of the cost of the surgical management. This compares well with other reports that also showed less financial cost of conservative approach compared to the surgical one(16–21). It helped in reducing the financial burdens on patients as well as the healthcare system. Our results showed that (8.18%) of the swapped patients in our cohort were confirmed to be COVID19 positive without classical respiratory symptoms like cough and fever highlighting the importance of our modified protocol that includes testing all patients admitted to our department during the COVID19 pandemic. In addition, our approach resulted in reduction in the number of patients presenting with acute abdomen, who tested positive for COVID19 and needed surgical intervention to only 1 patient (11.11%) out of the 9 COVID19 patients. This is essential in reducing the possible preoperative, intraoperative and postoperative viral transmission risks(14). Conservative approach is also essential in reducing the risk of surgery related complications in COVID19 patients who were found to suffer from higher mortality rates following different types of surgeries including minor procedures(22). This is attributed to their compromised lung functions, in addition to multiple organ dysfunction(12).

Interestingly, our protocol also included the implementation of telemedicine-based follow-up as a feasible and safe approach for follow up and evaluation of patient outcome. Such practices might be beneficial for both patients and healthcare providers through empowering social distancing to reduce the risk of viral transmission as well as reducing the pressure on the health care system.

In conclusion, our results showed that the implementation of conservative management in treating patients with acute appendicitis who were COVID19 positive is a safe and feasible approach that maybe essential in reducing preoperative, intraoperative and postoperative viral transmission risks as well as avoiding operative risks on COVID19 positive patients. Longer follow-ups to determine the true recurrence rate among the conservatively treated group as well as a larger number of patients might be needed in future studies.

## Data Availability

All data are included in this manuscript

